# Prevalence and factors associated with high-risk oncogenic human papillomavirus infection among women living with HIV in Kinshasa, the Democratic Republic of the Congo

**DOI:** 10.64898/2026.02.22.26346809

**Authors:** Julien Neze-Sebakunzi, Anna-Maria Doro Altan, Susanna Ceffa, Giovanni Guidotti, Stefano Capparucci, Fausto Ciccacci, Marlène Musikingala, Antoine Nkuba-Ndaye, Jean-Claude Makangara-Cigolo, Trésor Kabeya-Mampuela, Stefano Orlando, Steve Ahuka-Mundeke

## Abstract

**Background:** Cervical cancer is one of the most common cancers in women, particularly among women living with HIV (WLWH). Persistent infection with High-risk oncogenic human papillomavirus (Hr-HPV) is the primary etiological factor. However, data on Hr-HPV prevalence among WLWH in Kinshasa, Democratic Republic of the Congo, remains poorly documented. This study aimed to determine the prevalence of Hr-HPV infection and identify associated risk factors in this population.

**Methods:** A cross-sectional study was conducted on WLWH aged 25 to 65 years receiving antiretroviral therapy at the DREAM Centre in Kinshasa. Cervical sample were collected and analysing using multiplex PCR for detection of Hr-HPV genotypes. Sociodemographic data and risk factors were collected via questionnaires, and associations with Hr-HPV infection were assessed using multivariate logistic regression.

**Results:** A total of 436 women were included. The prevalence of Hr-HPV infection was 47.25%. HPV types 16 and 18 (alone or in co-infection) were detected in 23.79% of participants. In a multivariate logistic regression analysis, WHO clinical stage 3–4 (aOR 1.75; 95% CI 1.16–2.64; p=0.008) and HIV viral load ≥1000 copies/mL (aOR 3.08; 95% CI 1.28–7.42; p=0.012) and Antiretroviral therapy duration <2 years (aOR 0.52; 95% CI 0.29–0.93; p=0.028) were significantly associated with Hr-HPV infection.

**Conclusions:** Nearly one in two WLWH in Kinshasa was infected with Hr-HPV, and one in four carried HPV-16/18 genotypes. Advanced HIV disease and uncontrolled viral replication were strongly associated with Hr-HPV infection. These findings underscore the urgent need to integrate systematic Hr-HPV screening into HIV care programs, particularly for women with advanced clinical stage or persistent viremia.

## 1. Introduction

Cervical cancer (CC), classified as an AIDS-defining cancer (1,2), constitutes a major public health problem (3,4). It is one of the leading causes of cancer mortality worldwide and ranks as the fourth most common cancer in women (1,5–7). In 2020, approximately 604,000 women were diagnosed with CC, resulting in approximately 342,000 deaths (6). Nearly 90% of CCs occur in low- and middle-income countries (1,3,7,8), where it is often the leading or second leading cause of cancer and death in women. (7,9) In the Democratic Republic of the Congo (DRC), the incidence of CC is high, surpassing that of breast cancer (10,11).

The main causative agent of CC is the human papillomavirus (HPV) (3,6,7). Genital HPV infection is the most common sexually transmitted infection (STI) worldwide (12,13), with some researchers calling it the “sex flu” (12). Sexually active women are particularly at risk, and the persistence of this infection is critical for the development of CC (1,14). Approximately 200 HPV species have been identified (15), and are classified into low-risk (Lr-HPV) or high-risk (Hr-HPV) subtypes based on their oncogenic potential (8,11). Low-risk types can be asymptomatic or cause anogenital warts, while high-risk types are the cause of CC (3). Types 16 and 18 are responsible for approximately 70% of CC cases worldwide (1,16,17). Women living with HIV are at increased risk of precancerous lesions and cervical cancer compared to HIV-negative women (2,4,13,18). HIV-induced immunosuppression is associated with a higher prevalence of HPV, including high-risk types (9,19). HPV infections in HIV-positive women are not only more frequent, but they also persist more often, leading to an increased prevalence of high-grade cervical lesions (2,9). Unlike some opportunistic infections, such as Pneumocystis jiroveci pneumonia, extrapulmonary cryptococcosis, or histoplasmosis, whose incidence has decreased after the introduction of antiretroviral therapy, the incidence of CC has not decreased significantly. This is because persistent immunosuppression, a characteristic feature of people living with HIV, remains a major risk factor for the development of virally related cancers, including HPV (3). The risk of developing cervical cancer is six times higher in women living with HIV than in those who are not infected (17,20).

In high-income countries, cervical cancer incidence and mortality have fallen by more than 50% over the past three decades, thanks to the implementation of national screening programs. On the other hand, in low- and middle-income countries, the absence or weakness of screening and vaccination initiatives against human papillomavirus (HPV) continues to contribute to the persistence of this public health problem (3).

Cervical cancer is largely preventable through vaccination and screening for precursor lesions, with appropriate follow-up and treatment (4). Early screening of women at risk of developing cervical lesions is essential to reduce the incidence of cervical cancer (6). As part of the second edition of the World Health Organisation (WHO) guidelines, published in July 2021 for the screening and treatment of precancerous cervical lesions, it is now recommended to prioritize HPV deoxyribonucleic acid (DNA) detection as the primary screening test, replacing visual inspection with acetic acid (VIA) or cytology, both for the general population and for women living with HIV (17). Indeed, recent research has explored the use of HPV testing as a primary screening modality, demonstrating that it is as effective, or even more than cytology alone (8,17).

However, although a 2019 study reported a prevalence of 24.4% of Hr-HPV infections in the general female population of Kinshasa (3), there is a lack of specific data regarding women living with HIV in this region.

Therefore, our study aims to estimate the prevalence and distribution of Hr-HPV infections among women living with HIV in Kinshasa, while identifying the factors associated with these infections.

## 2. Methods

### 2.1. Study Type, Setting, and Population

We conducted an observational cross-sectional study from April 2022 to April 2023. The study population included all women aged 25 to 65 years receiving antiretroviral therapy (ART) at the DREAM Centre in Kinshasa, a facility specialized in the care of people living with HIV (PLWH). Women were asked to participate and were enrolled progressively, based on their attendance at scheduled medical check-up appointments during the study period. To limit non-inclusion bias, all women who missed their appointments received phone call reminders to encourage them to attend their appointments and participate in the screening.

Written informed consent was obtained from all participants. Pregnant women were excluded as a precaution to avoid any potential risk or even subjective discomfort related to cervical sampling. Women who were menstruating at the time of sample collection were rescheduled to ensure standardized sampling conditions and were invited to return at least one week after the end of their menstrual period. Women with a history of hysterectomy, prior HPV testing, or previous HPV vaccination were also excluded.

The required sample size (n) is at least 385 participants. It was calculated assuming a prevalence of 50% due to the absence of precise data, a 95% confidence interval (with a Z-score of 1.96), and a precision of 5%:

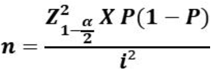

### 2.2. Data collection and analysis

Data were collected using a structured and standardized questionnaire, including demographic (age, provenance, education, employment status, marital status), reproductive (menopause, contraceptive use, age at first intercourse, abortion history, parity, number of lifetime sexual partners), and clinical information (weight, height for Body mass index (BMI)). For the purposes of this study, the variable “provenance” was categorized into two groups : “intra-zonale” and “extra-zonale.” “Intrazonal” refers to participants residing within the municipality where the health facility is located (the municipality of N’Sele), while “extrazonal” refers to participants coming from outside this municipality. It is important to note that the municipality of N’Sele is considered an urban–rural area of Kinshasa. Additionally, WHO clinical stage and ART duration were extracted from patients’ electronic medical records.

For HIV viral load (VL), venous blood was collected in Ethylene Diamine Tetraacetic Acid (EDTA) tubes, plasma separated by centrifugation, and stored at −20°C. VL was measured using Abbott RealTime HIV-1 on the m2000 platform (Abbott Molecular, Abbott Park, IL, USA), following manufacturer’s protocols. RNA was extracted from 1 ml of plasma using magnetic particle technology and amplified by PCR (21).

Cervical sample were collected using the Abbott Cervi-Collect™ Specimen Collection Kit (Abbott Molecular, Wiesbaden, Germany) by a trained physician. After insertion of a vaginal speculum and identification of the squamocolumnar junction, the Cervi-Collect brush was gently inserted into the cervical canal and rotated three times in the same direction to collect epithelial cells. The brush was then immediately placed into the Cervi-Collect transport tube containing the proprietary transport medium. Samples were then stored at 2°C to 8 °C for up to two weeks at the DREAM Laboratory in Kinshasa to standardize storage conditions across all samples, in accordance with the manufacturer’s instructions, which allow storage between 2 °C and 30 °C for up to 14 days.

Samples were analyzed using the Abbott RealTime High Risk HPV assay on the m2000 platform (Abbott Molecular, USA), according to the manufacturer’s instructions. This qualitative real-time PCR assay detects DNA from 14 high-risk HPV genotypes, with individual identification of HPV-16 and HPV-18 and pooled detection of 12 other high-risk types (HPV-31, -33, -35, -39, -45, -51, - 52, -56, -58, -59, -66, and -68). The analytical performance of this assay has been previously described (23).

Data processing and analysis were performed using Stata® 14 (StataCorp, 2014). The prevalence of Hr-HPV was estimated with 95% confidence intervals (CI). Categorical variables were presented as proportions and continuous variables were summarized as mean ± standard deviation (SD) for normally distributed data, or as median and interquartile range (IQR) for skewed distributions. The normality of the data was assessed both visually, using density curves overlaid on histograms, and statistically, using the Shapiro–Wilk test. Univariate and multivariate logistic regression models were used to identify factors associated with Hr-HPV among people living with HIV/AIDS. Variables with p < 0.2 in univariate analysis or known relevance based on existing literature were included in the multivariate logistic regression model. Adjusted odds ratios (aOR) with 95% CI were calculated to assess associations with Hr-HPV infection; p < 0.05 was considered significant.

### 2.3. Ethical considerations

This study was conducted in accordance with the Declaration of Helsinki. Its conduct was approved by the Ethics Committee of the School of Public Health of the University of Kinshasa, under approval number ESP/CE/31/2022, dated April 27, 2022.

All participants gave their written informed consent and had the option to withdraw from the study at any time, without any negative consequences.

The data collected was treated with the strictest confidentiality and anonymity. Participants were reassured to receive their test results free of charge, and those whose results were positive were referred to the care unit where they received additional assessments and appropriate treatment.

## 3. Results

### 3.1. Study Participant Flow

Of the 524 HIV-positive women aged 25 to 65 years and receiving antiretroviral therapy, 451 were enrolled. The remaining 73 were not enrolled for the following reasons: 23 women were pregnant, 36 were menstruating, 1 woman had a history of hysterectomy, and 13 refused consent. Of the 451 enrolled, 15 women were excluded: 11 for refusal to sample (due to fear or lack of preparation) and 4 others for bleeding observed during the attempted cervical sampling. Thus, 436 women were included in the study (Figure 1).

**Figure. 1.**
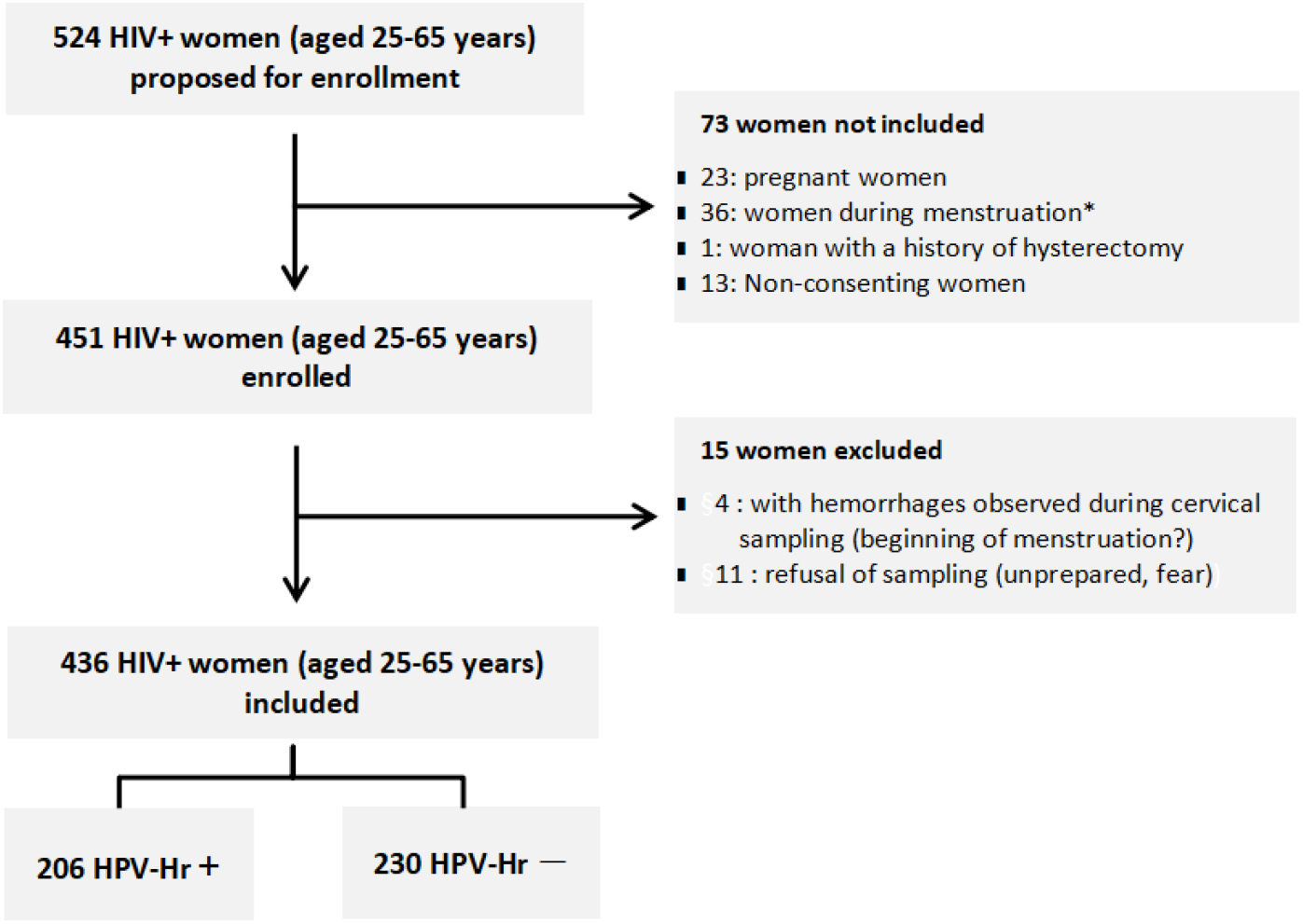
Participant flow diagram. **Note** : * Women who were menstruating at the time of sample collection were temporarily excluded and invited to return for testing at least one week after the end of their menstrual period, in order to standardize the timing of sample collection. Those who did not return despite follow-up reminders were excluded.

### 3.2. Demographic, Reproductive, Clinical, and Biological Characteristics

The mean age of participants was 44.46 years (± 9.02). Women aged 25 to 29 had the highest positivity rate for Hr-HPV (60%), while older age groups showed a decrease, reaching 43.79% among those aged 40 to 49. Women living outside the N’sele health zone had a slightly lower positivity rate (45.54%) than those living in this zone (49.06%). The rate was lowest among those with a university degree (37.50%). Employed women and women in a relationship had lower positivity rates (39.02% and 40.11%) than those unemployed or single. (Table 1)

**Tab 1.**
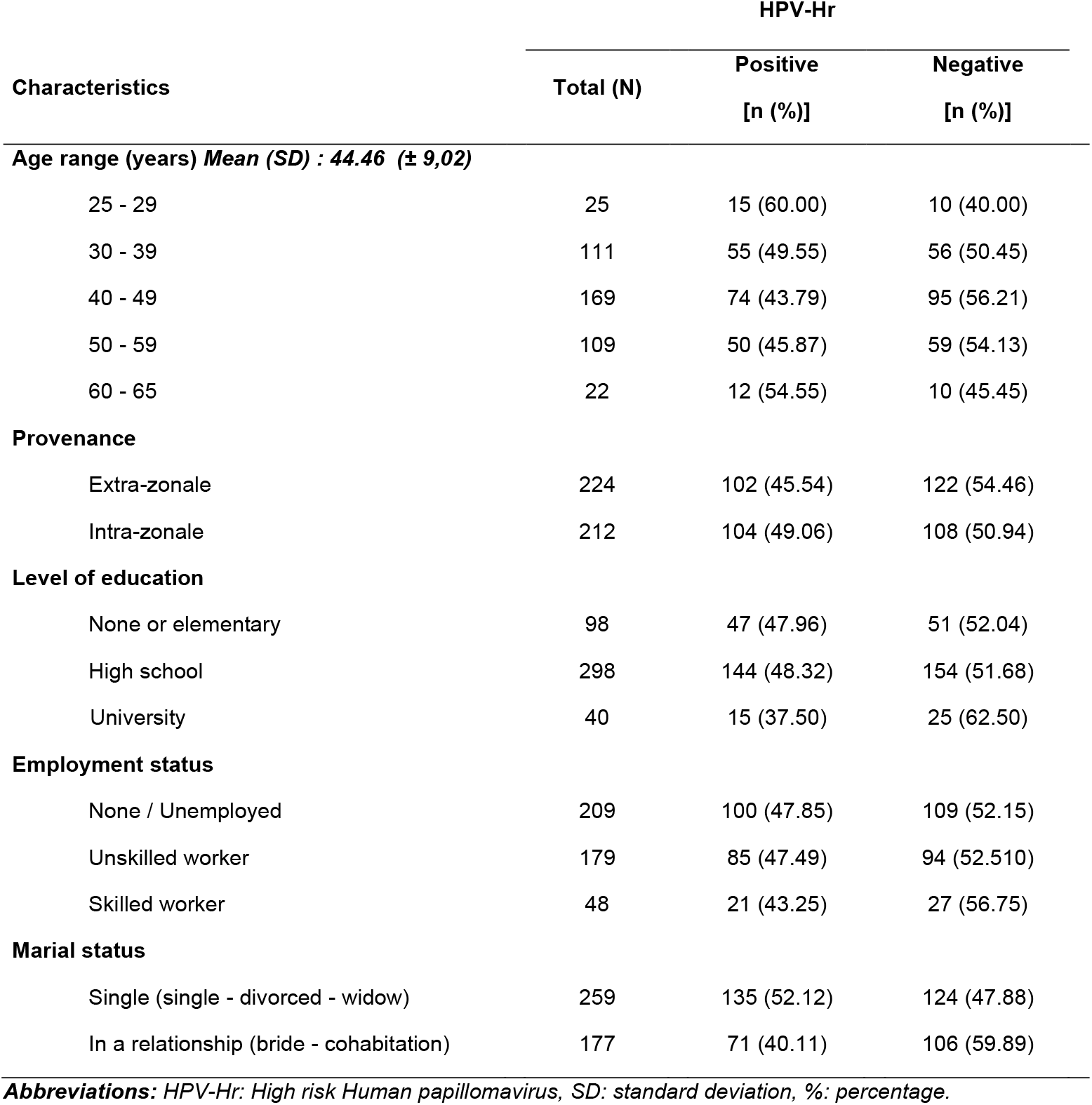
Sociodemographic characteristics and Hr-HPV screening results among women living with HIV, aged 25 to 65, receiving antiretroviral therapy at the DREAM Center in Kinshasa, Democratic Republic of Congo [n = 436].

| Characteristics | HPV-Hr |  |  |
| --- | --- | --- | --- |
|  | Total (N) | Positive | Negative |
|  |  | [n (%)] | [n (%)] |
| Age range (years) Mean (SD) : 44.46 (± 9,02) |  |  |  |
| 25 - 29 | 25 | 15 (60.00) | 10 (40.00) |
| 30 - 39 | 111 | 55 (49.55) | 56 (50.45) |
| 40 - 49 | 169 | 74 (43.79) | 95 (56.21) |
| 50 - 59 | 109 | 50 (45.87) | 59 (54.13) |
| 60 - 65 | 22 | 12 (54.55) | 10 (45.45) |
| Provenance |  |  |  |
| Extra-zonale | 224 | 102 (45.54) | 122 (54.46) |
| Intra-zonale | 212 | 104 (49.06) | 108 (50.94) |
| Level of education |  |  |  |
| None or elementary | 98 | 47 (47.96) | 51 (52.04) |
| High school | 298 | 144 (48.32) | 154 (51.68) |
| University | 40 | 15 (37.50) | 25 (62.50) |
| Employment status |  |  |  |
| None / Unemployed | 209 | 100 (47.85) | 109 (52.15) |
| Unskilled worker | 179 | 85 (47.49) | 94 (52.510) |
| Skilled worker | 48 | 21 (43.25) | 27 (56.75) |
| Marial status |  |  |  |
| Single (single - divorced - widow) | 259 | 135 (52.12) | 124 (47.88) |
| In a relationship (bride - cohabitation) | 177 | 71 (40.11) | 106 (59.89) |

**Abbreviations:** HPV-Hr: High risk Human papillomavirus, SD: standard deviation, %: percentage.

Having first sexual intercourse at age 15 or younger was associated with a positivity rate of 51.41%. Women with HIV stages 3 and 4 had a 55.07% HPV-HR positivity rate, while those with an HIV viral load greater than or equal to 1000 copies/mL had the highest rate (71.43%). Women on antiretroviral therapy for 2 years or less had a positivity rate of 61.54%. (Table 2)

**Tab 2.** Clinical, reproductive, and laboratory characteristics and HR-HPV screening results among women living with HIV, aged 25 to 65 years, receiving antiretroviral therapy at the DREAM Center in Kinshasa, DRC [n = 436].

| Characteristics | HPV-Hr |  |  |
| --- | --- | --- | --- |
|  | Total (N) | Positive [n (%)] | Negative [n (%)] |
| **Menopause** |  |  |  |
| No | 253 | 121 (47.83) | 132 (52.17) |
| Yes | 183 | 85 (46.45) | 98 (53.55) |
| **Contraceptive method** |  |  |  |
| None | 384 | 181 (47.14) | 203 (52.86) |
| Hormonal | 52 | 25 (51.92) | 27 (48.08) |
| **Age at first sexual intercourse (years) Median (IQR) : 16.51 (15 - 18)** |  |  |  |
| ≤15 | 142 | 73 (51.41) | 69 (48.59) |
| 16 - 20 | 272 | 127 (46.69) | 145 (53.31) |
| ≥ 21 | 22 | 10 (45.45) | 12 (54.55) |
| **Number of sexual partners (lifetime)** |  |  |  |
| < 4 | 185 | 84 (45.41) | 101 (54.59) |
| 4 – 7 | 154 | 70 (45.45) | 84 (54.55) |
| ≥ 8 | 97 | 52 (53.61) | 45 (46.39) |
| **History of abortion Median (IQR) : 1.32 (0 - 2)** |  |  |  |
| No | 159 | 82 (51.57) | 77 (48.43) |
| Yes | 277 | 124 (44.77) | 153 (55.23) |
| **Parity Median (IQR) : 3.87 (2 - 4)** |  |  |  |
| ≤ 4 | 258 | 116 (44.96) | 142 (55.04) |
| > 4 | 178 | 90 (50.56) | 88 (49.44) |
| **Body mass index (Kg/m<sup>2</sup>) Median (IQR) : 24.36 (20.55 – 27.60)** |  |  |  |
| <18.5 | 49 | 31 (63.27) | 18 (36.73) |
| 18.5 - <25 | 218 | 104 (47.71) | 114 (52.29) |
| ≥ 25 - <30 | 112 | 48 (42.86) | 64 (57.14) |
| > 30 | 57 | 23 (40.35) | 34 (59.65) |
| **WHO Clinical Stage (HIV)** |  |  |  |
| 1 & 2 | 209 | 81 (38.76) | 128 (61.24) |
| 3 & 4 | 227 | 125 (55.07) | 102 (44.93) |
| **HIV Viral load (copies/mL)** |  |  |  |
| < 40 | 358 | 159 (44.41) | 199 (55.59) |
| 40 - < 1000 | 50 | 27 (54.00) | 23 (46.00) |
| ≥ 1000 | 28 | 20 (71.43) | 8 (28.57) |
| **Duration of ART (years)** |  |  |  |
| < 2 ans | 65 | 40 (61.54) | 25 (38.46) |
| ≥ 2 ans | 371 | 166 (44.74) | 205 (55.26) |

**Abbreviations:** ART: antiretroviral therapy, HIV: Human Immunodeficiency Virus, HPV-Hr: High risk Human papillomavirus, IQR: Interquartile Range, Kg: kilogram, mL: milliliter, m<sup>2</sup>: square meter, WHO: World Health Organization, %: percentage.

### 3.3. Prevalence of Hr-HPV infection and distribution of Hr-HPV genotypes

The prevalence of Hr-HPV infection in the study population was 47.25% (206/436; 95% CI: 42.58–51.96) (Table 3).

**Tab 3.** Prevalence of Hr-HPV among women living with HIV, aged 25–65 years, receiving antiretroviral therapy at the DREAM Center in Kinshasa, DRC [n = 436].

| Hr-HPV | Frequency | % (CI 95%) |
| --- | --- | --- |
| Negative | 230 | 52.75 (48.04 – 57.42) |
| Positive | 206 | 47.25 (42.58 – 51.96) |

**Abbreviations:** CI: Confidence interval, Hr-HPV: High risk Human papillomavirus, %: percentage.

Regarding genotype distribution, the majority of Hr-HPV infections (76.21%; 95% CI : 69.87– 81.56) were attributable to high-risk genotypes other than HPV-16 and HPV-18. Overall, HPV-16 and/or HPV-18 (alone or in co-infection) were detected in approximately one quarter of Hr-HPV-positive women.

HPV-16 was identified in 6.8% of participants (95% CI : 4.05–11.19), and HPV-18 in 5.34% (95% CI : 2.97–9.43). Coinfections involving HPV-16 or HPV-18 were less frequent, and simultaneous detection of HPV-16, HPV-18, and other genotypes was rare (0.49%; 95% CI : 0.06– 3.42) (Figure 2).

**Figure. 2.**
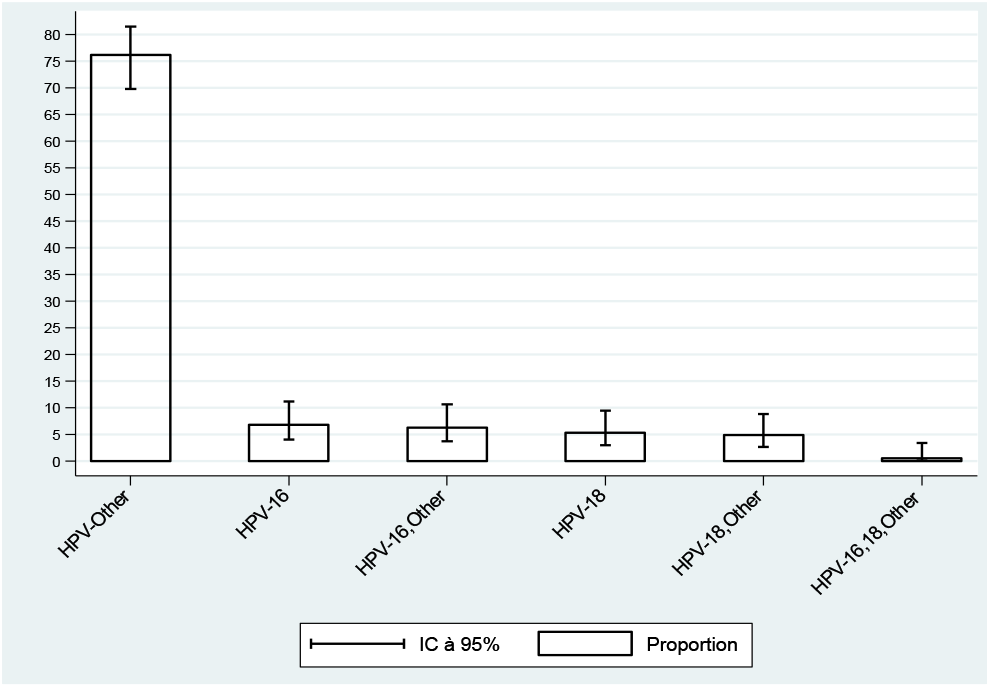
Hr-HPV genotype distribution among women living with HIV, aged 25–65 years, under antiretroviral treatment at the DREAM Center in Kinshasa, DRC [n = 206]

### 3.4. Associated Factors

In the univariate analysis, several variables such as marital status, body mass index, HIV clinical stage, duration of antiretroviral therapy, and viral load, showed a significant association with Hr-HPV infection (Tab 4).

**Tab 4.**
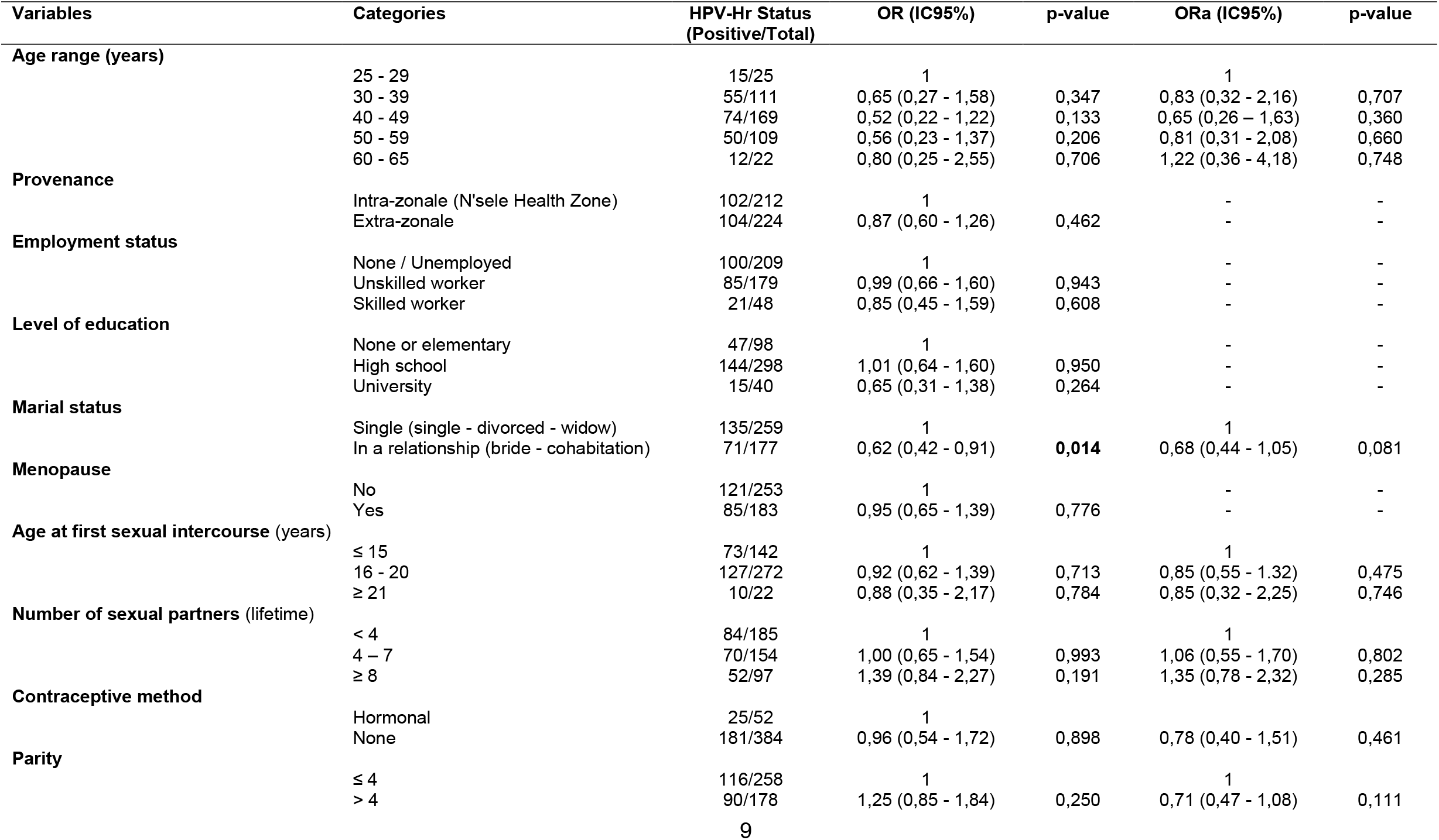
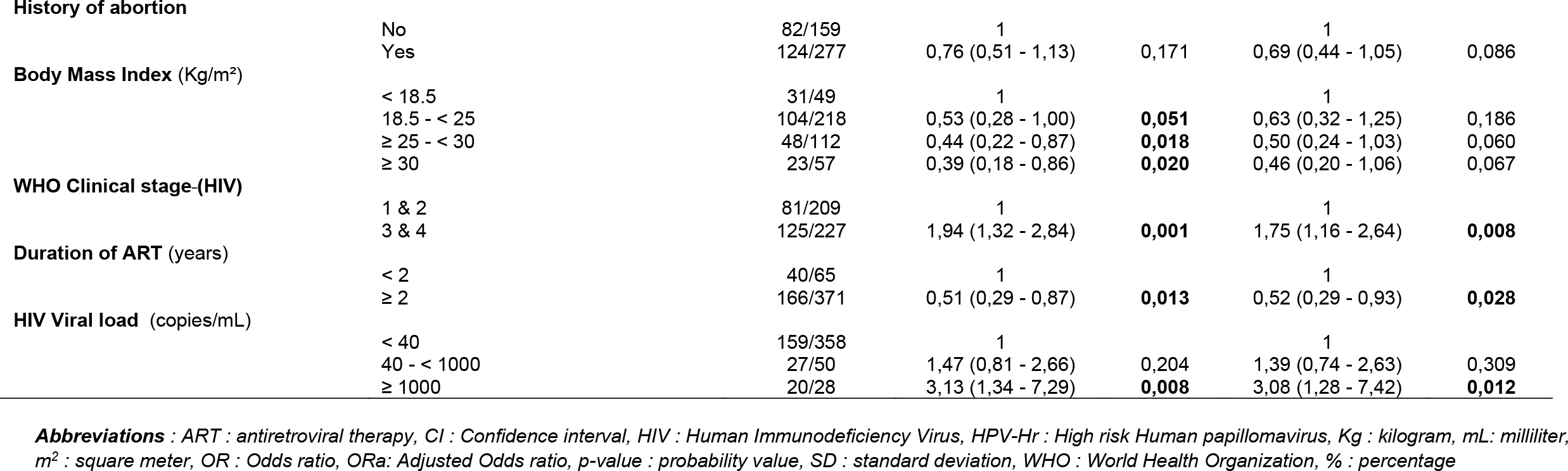
Associations between socio-demographic and clinical characteristics and HPV-Hr status in women living with HIV, aged 25 to 65 years, receiving antiretroviral treatment at the DREAM Center in Kinshasa, DRC [n = 436].

In a multivariate analysis, only the association with HIV clinical stage, duration of antiretroviral therapy, and viral load remained statistically significative.

The results suggest that women with advanced HIV or high viral load above 1000 copies/ml are more likely to be infected with Hr-HPV, and that prolonged antiretroviral therapy may play a protective role.

## 4. Discussion

This study was conducted to describe the characteristics of Hr-HPV in women living with HIV in the city-province of Kinshasa, DRC. We estimated the prevalence and distribution of Hr-HPV infections and identified associated factors.

The results showed a particularly high prevalence of Hr-HPV in cervical samples collected using a cytobrush, with an overall frequency of 47.25% among women living with HIV in Kinshasa. This prevalence is two to four times higher than those found in the general population in Kinshasa, which range from 12.5% to 24.4% (11,24,25). This discrepancy may be explained by the fact that our study only included HIV-positive women. Several studies suggest, in fact, that HIV infection constitutes an independent risk factor for Hr-HPV infection (6,18,20,26,27). Globally, approximately 1 in 20 CCs is attributable to HIV; in Sub-Saharan Africa, approximately 1 in 5 CCs is due to HIV (28). The prevalence of Hr-HPV found in this study was, however, higher than that found in other studies in Africa among women living with HIV (29–34). It is, however, lower than that found in another study in Burkina Faso and South Africa (35)-Prevalence on the continent varies by region and is heterogeneous among the populations studied (1,22). The large differences in incidence between different countries have also been influenced by the introduction of screening (1).

The results of this study also show that HPV-16 and HPV-18 types are present in approximately one quarter of all high-risk HPV positive participants, with nearly half of these infections corresponding to coinfections (11.65%), while the other half consists of isolated infections with HPV-16 or HPV-18 (12.14%). Furthermore, a significant proportion of infections (76.21%) were associated with Hr-HPV other than types 16 and 18. These findings corroborate the results of other studies where a large proportion of infections were found to be related to Hr-HPV other than types 16 and 18.(29,31–33) This observation is not surprising, as some studies in Africa have revealed distinct genotypic profiles, with types other than HPV-16 and HPV-18 predominating in certain populations at increased risk, such as women living with HIV (9,35). Hence, the need for a more comprehensive approach in CC prevention strategies, as well as the importance of conducting preliminary studies to identify the most common HPV types and adapting vaccines accordingly. The high prevalence of HPV-35 among women with confirmed CC in Zimbabwe provides further evidence of the diversity of HPV genotypes in sub-Saharan Africa (36). Thus, women living with HIV in sub-Saharan Africa are less likely to be covered by the available vaccine, even the nine-valent, mainly due to the lower prevalence of HPV16 and the higher prevalence of HPV35 (37). In this context, it becomes essential to offer vaccination covering a wider range of HPV genotypes and to use more comprehensive screening tests, in line with the objectives of the WHO 90-70-90 strategy for the elimination of this cancer, where 90% of girls should be vaccinated before the age of 15, and 70% of women - age - should be screened (6).

The results of this study further suggest that the clinical stage of HIV, the duration of treatment antiretroviral therapy and HIV viral load play a key role in HPV prevalence among women living with HIV in Kinshasa.

Women with HIV stages 3 and 4 have a significantly higher risk of HPV infection. Our results are similar to previous studies, which have reported that as the clinical condition of people living with HIV deteriorates, their immune system weakens, increasing their risk of HPV infection (29,38).

Also, women who have been on ARV treatment for more than 2 years have a reduced risk of HPV prevalence. These results corroborate those found in a study conducted in Burkina Faso and South Africa where people taking ARVs for less than 2 years had a higher risk (35). Another study in Ghana identified highly active antiretroviral therapy as a protective factor against Hr-HPV (39). However, it should be noted that the effect of antiretroviral treatment on cervical neoplasia is little known and is still the subject of debate, although ART in HIV-infected women has a clear effect on restoring immune status (9).

Furthermore, in this study, women with a VL ≥ 1000 copies/mL were approximately three times more likely to be infected with Hr-HPV than those with a VL < 40 copies/mL. This finding is supported by several recent studies, including a systematic review and a meta-analysis, which highlight the importance of antiretroviral therapy (ART), CD4 count, and viral load in the risk of HPV infection and the development of cervical cancer (40). Furthermore, a study in pregnant women showed that those who achieved viral suppression were less likely to acquire Hr-HPV infection (41), highlighting the benefits of treatment adherence and viral suppression in reducing the risk of progression to HPV-related pathologies (38,40–42).

## 5. Limitations

Although this study provides important information to guide our health policies, it has certain limitations. First, the monitoring of patients on ART was primarily focused on measuring HIV viral load, and CD4 counts were not included. Inclusion of CD4 data would have allowed a more precise evaluation of the relationship between immunosuppression and Hr-HPV infection.

Furthermore, the 12 high-risk HPV genotypes other than HPV-16 and HPV-18 were analyzed as a pooled group, preventing genotype-specific assessment and limiting detailed analysis of their individual distribution. Such information could be relevant for optimizing prevention strategies, including vaccine implementation.

Finally, due to the cross-sectional design of the study, causal relationships between associated factors and Hr-HPV infection cannot be established.

## 6. Conclusion

This study revealed a high prevalence of Hr-HPV infections among women living with HIV followed at the DREAM Center in Kinshasa, with nearly one in two women affected and one in four carrying HPV-16/18 genotypes. Advanced HIV disease and uncontrolled HIV viral replication or less than two years of antiretroviral therapy are at increased risk of carrying Hr-HPV.

These findings highlight the need to integrate routine Hr-HPV screening into HIV care programs to enable early detection and appropriate management. Strengthening viral load control and clinical follow-up among women with advanced HIV disease may contribute to reducing the burden of HPV co-infection.

In the absence of HPV vaccination in the national immunization program of the DRC, the introduction of HPV vaccination should be considered as part of a comprehensive strategy to prevent cervical cancer and support global elimination efforts.

## Data Availability

All data generated or analyzed during the current study are included within the manuscript. Additional data are available from the corresponding author on reasonable request.

## Abbreviations

AIDS: Acquired Immunodeficiency Syndrome
aOR: Adjusted Odds Ratio
ART: Antiretroviral Therapy
CC: Cervical Cancer
DRC: Democratic Republic of the Congo
HIV: Human Immunodeficiency Virus
HPV: Human Papilloma Virus
Hr-HPV: High-Risk Human Papilloma Virus
PLWH: People Living With HIV
VL: Viral Load
WLWH: Women Living With HIV
WHO: World Health Organization

## Ethics approval and consent to participate

This study was conducted in accordance with the principles of the Declaration of Helsinki. Its conduct was approved by the Ethics Committee of the School of Public Health of the University of Kinshasa, under approval number ESP/CE/31/2022. All participants provided written informed consent and had the option to withdraw from the study at any time, without any negative consequences. The data collected was treated with the strictest confidentiality and anonymity. Participants were reassured that they would receive their test results free of charge, and those who tested positive were referred to the care unit where they received additional assessments and appropriate treatment.

## Consent for publication

Not applicable

## Author contributions

Conception, design and supervision of the study: JNS, AMD, GG, SC, SAM; Acquisition of data: JNS, MM, SC, SCa; Analysis and interpretation of data: JNS, AMD, JCMC, SC, SCa, GG, TKM, ANN, FC, SO, SAM; Writing the first draft: JNS, AMD; Reviewed the first draft: JNS, AMD, SC, SCa, GG, JCMC, FC, SO, SAM; Revision of the submitted article: JNS, AMD, SC, SCa, GG, JCMC, MM, ANN, TKM, FC, SO, SAM.

## Acknowledgements

We thank the patients, nurses and other DREAM program staff in Kinshasa (DRCongo) who made this study possible. We also acknowledge Abbott for donating materials and reagents to perform the HPV test.

